# Neurophysiological Monitoring of Sport Related Concussion: From Diagnosis to Return to Play

**DOI:** 10.64898/2026.10.12.26365262

**Authors:** Jin Lee, Eric D. Kirby, Christopher Levi, Louis Shidiak, Andrew J. Gardner, Ryan C.N. D’Arcy

## Abstract

The validation of objective neurophysiological monitoring of concussion from diagnosis to return to play has the potential to improve the clinical care of athletes. Extracted from event-related potentials (ERP), ‘brain vital signs’ offer a non-invasive objective neurophysiological measure of three well established ERP responses that complement existing clinical assessments: 1) the N100 response related to auditory sensation; 2) the P300 response related to basic attention; and 3) the N400 response related to cognitive processing. The N400, in particular, has increasingly been demonstrated to be sensitive to head impact related cognitive impairment.

Accordingly, brain vital sign measures were collected in *n* =165 rugby league players from two Australian professional National Rugby League clubs (NRL), with concussed players (*n*=26) being subsequently assessed over a total of five separate time points (Baseline, Day 2, Day 5, Day 10, and Day 42). This timeline aligned with acute (Day 2), NRL mandatory minimum return-to-play (RTP) guidelines (Day 10), and an extended post RTP period.

Relative to baseline, the results revealed significant N400 cognitive processing latency delays at Day 2 post-concussion (p=0.002). The N400 latency delays persisted past day 5, significantly returning to baseline values at Day 10, consistent with the RTP mandatory minimum requirement. The findings indicated that neurophysiological recovery occurred in this cohort within an approximate 10-day time frame. Future studies monitoring brain vital sign changes across the trajectory of injury to recovery are needed in sports related concussion management.

## Introduction

### Background

Despite ongoing advancements in concussion care, determining recovery under the current clinical assessment framework relies on subjective clinical judgement and expert opinion. Given the recognised limitations of clinical assessment tools,^1,2^ there is increasing interest in objective measures such as neurophysiological markers to help inform subjective measures, principally symptom checklists.^3–9^ This presents a particular challenge in sports related concussion management in professional rugby league, where athletes in both the National Rugby League (NRL) and National Rugby League Women (NRLW) premierships have been noted to under-report or fail to disclose concussion symptoms.^10,11^ Further to this, clinical tests, such as the SCAT6, have been reported to have limited utility when administered > 7 days post-concussion.^3^

The latest NRL concussion protocol^12^ requires SCAT6^13^ assessment and Cognigram^TM14^ testing when determining recovery and clearing an athlete to return to play (RTP). In addition to these tests, athletes are required to progress through a graduated protocol in line with the latest Concussion in Sport consensus statement^7^ in which athletes are prescribed increasing physical and cognitive loads whilst remaining symptom free. There remains, however, considerable variation in RTP timeframes and measures used across clinical practice when determining RTP readiness.^15–17^ Consequently, biomarkers such as neurophysiological measures have potential to guide clinical decision making and advance post-concussion monitoring over multiple time points across the trajectory of recovery.

### Brain vital signs

Event related potentials (ERPs) have emerged as a promising objective evaluation of cognitive brain function changes following concussion.^18–20^ Advances in electroencephalography (EEG) and ERP applications have led to the emergence of a ‘brain vital signs’ framework, translating ERPs into a clinically accessible, point-of-care measure of cognitive brain function.^20–23^

Brain vital signs incorporate standardised amplitudes and latency metrics of well-established ERP responses: the N100 related to auditory sensation; the P300 related to basic attention; and the N400 related to cognitive processing.^20,21^ Importantly, a number of studies have utilized brain vital signs as an objective neurophysiological measure in concussion and non-concussive repeated head impacts (RHI) across a range of sporting populations.^24–26^ The N400 response, in particular, has demonstrated repeatable sensitivity to RHIs, significantly detectable across studies as changes in either peak latency or amplitude, or both.^27,28^

One of the most pressing questions emerging from neurophysiological studies relates to characterising the trajectory of ERP change from diagnosis to recovery, during the time frame to RTP. This central question focuses directly on the potential for brain vital signs to be used as a clinical decision assistance tool in determining cognitive recovery and readiness for a concussed athlete to return to play.

### Objective and hypotheses

The objective of this study was to investigate brain vital signs as an objective neurophysiological measure in determining cognitive recovery following concussion. The primary hypothesis of the study was that the N100, P300 and N400 responses would be sensitive to concussion and subsequently normalise (i.e., return to individual baseline levels) in the time points following concussion.

## Material and Methods

### Design

This study was an observational, prospective within-subject cohort study. Organisational consent and support for the study was provided by the two participating NRL clubs and their management. Athletes provided informed written consent in accordance with the Declaration of Helsinki and approved by the ethical committee of Hunter New England Health [2023/ETH01645].

### Participants

Both male and female professional rugby league athletes undertook baseline brain vital sign assessments during the 2024 and 2025 NRL/NRLW premiership seasons across two NRL clubs (four teams total). Athletes subsequently medically diagnosed with concussion became eligible for continued follow up using ERP measures. Athletes were medically diagnosed with concussion, informed in part by video evidence on game day/training and by clinical assessment as per the NRL’s concussion protocol.^12^ Exclusion criteria included athletes less than 18 years of age or greater than 35 years of age, not proficient in the English language, clinically documented issues with hearing, and allergies to rubbing alcohol or EEG gel. There were no athletes excluded from the study based off the above criteria.

## Measures and methods

### Brain vital sign ERPs

Brain vital sign ERPs were collected with the NeuroCatch**^®^** Platform. NeuroCatch**^®^** is a point of care medical device with Class IIa regulatory approval from the Therapeutic Goods Administration (ARTG ID 439393, Sponsor CXDX Pty Ltd). NeuroCatch**^®^** automatically extracts standardized amplitude and latency measures for three brain vital sign ERPs: 1) The N100^29^ is representative of auditory sensation; 2) the P300,^30^ a measure of basic attention; and 3) N400^31^, a measure of cognitive processing. Individual brain vital sign results are provided relative to normative data for all six metrics (N100, P300, N400; amplitude and latency) using a standardized radar plot format, along with the recorded ERP responses.

### Data Collection

Athletes underwent baseline NeuroCatch**^®^** assessment at the commencement of the 2024 and 2025 NRL and NRLW seasons. In addition, routine SCAT6^13^ and Cognigram^TM^ pre-season evaluations were carried out as part of the NRL’s baseline testing framework.^12^

The NRL/NRLW’s concussion protocol^12^ followed the latest Consensus statement on concussion in sport^7^. In brief, this protocol requires a minimum 11 day return to play timeframe and mandated medical follow to monitor clinical recovery prior to returning to play. Progression to the final stages of protocol, including the re-introduction of contact training, is contingent upon normalisation of SCAT6 and post-concussion Cognigram^TM^ scores to the individual’s pre-season baseline and progression through progressive aerobic loads whilst remaining symptom free.

Following concussion diagnosis, brain vital sign monitoring used a total of five timepoints for each athlete (Baseline, Day 2, Day 5, Day 10, and Day 42). NeuroCatch**^®^** scans were carried out in parallel, but independent of, the existing clinical protocol (Fig. 1). Day 2, the second time point coincided with the initial follow up appointment, undertaken to monitor recovery following concussion and to identify athletes with delayed onset of concussion symptoms. Day 5, the third time point was selected at approximately 5 days post injury. This time point was selected to reflect a common time point when athletes had completed stage 3 of the graded return to play protocol and were undertaking stage 4 of the protocol. Day 10, the fourth time point was selected just ahead of the mandatory minimum RTP requirement. Finally, Day 42, the fifth time point was selected approximately 42 days (6 weeks) post injury, reflective of a subacute recovery timeframe following concussion.

**Figure 1:**
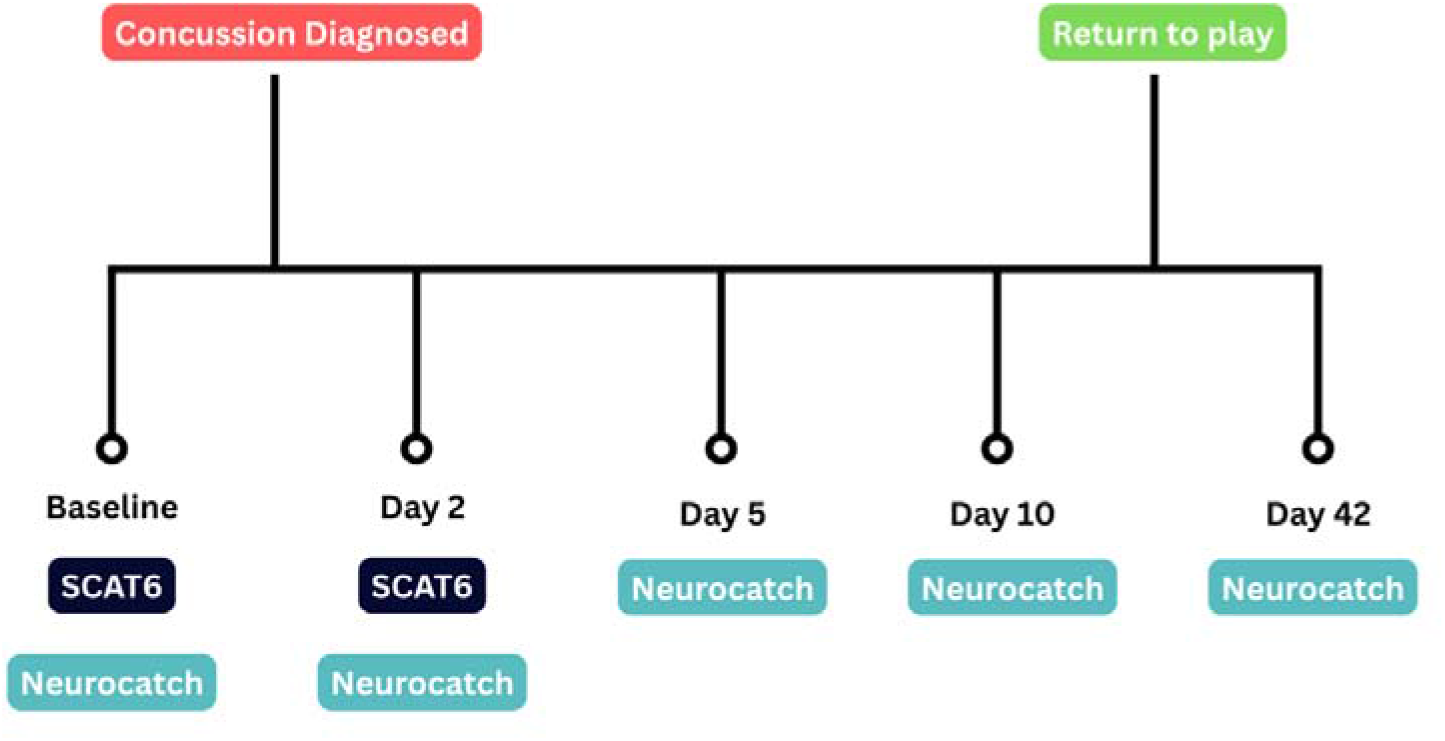
Study flowchart. Athletes followed the NRL’s concussion RTP protocol and NeuroCatch**^®^**scans were performed on five separate occasions.

All tests were completed by a member of the wider medical staff at the respective rugby league clubs, who were not involved in making clinical decisions around return to play. NeuroCatch**^®^** data analyses followed a two-step verification using standardized report measures compared against both general normative data and NRL specific normative data using the *n*=165 baseline data.

### Clinical recovery and RTP decision making

RTP decisions at both clubs were made solely on clinical grounds in accordance with the NRL/NRLW protocol.^12^

### Statistical analysis

Brain vital sign ERP analyses were based on the group average radar plot findings (Figure 2), to evaluate statistical differences between baseline and Day 2 along with any other differences in the recovery trajectory for Day 5, Day 10, and Day 42. Results are presented as mean (+/-Standard Error bars). Time point results were compared using a linear mixed model (LMM) that treated each brain vital sign as the dependent measure (six separate LMMs: N100, P300, and N400 amplitude and latency) and included timepoint (5 levels: baseline, 2-day post concussion, 5-day post concussion, and 42 day post concussion) as the single fixed effect. Each participant was treated as a random effect to account for individual change trajectories. Models were fit using restricted maximum likelihood (REML). As a post-hoc analysis of individual timepoints, pairwise comparisons of estimated marginal means across timepoints were conducted for brain vital signs with significant main effects of timepoint. Each pairwise comparison was adjusted for multiple comparisons with Bonferroni adjustment.

**Figure 2:**
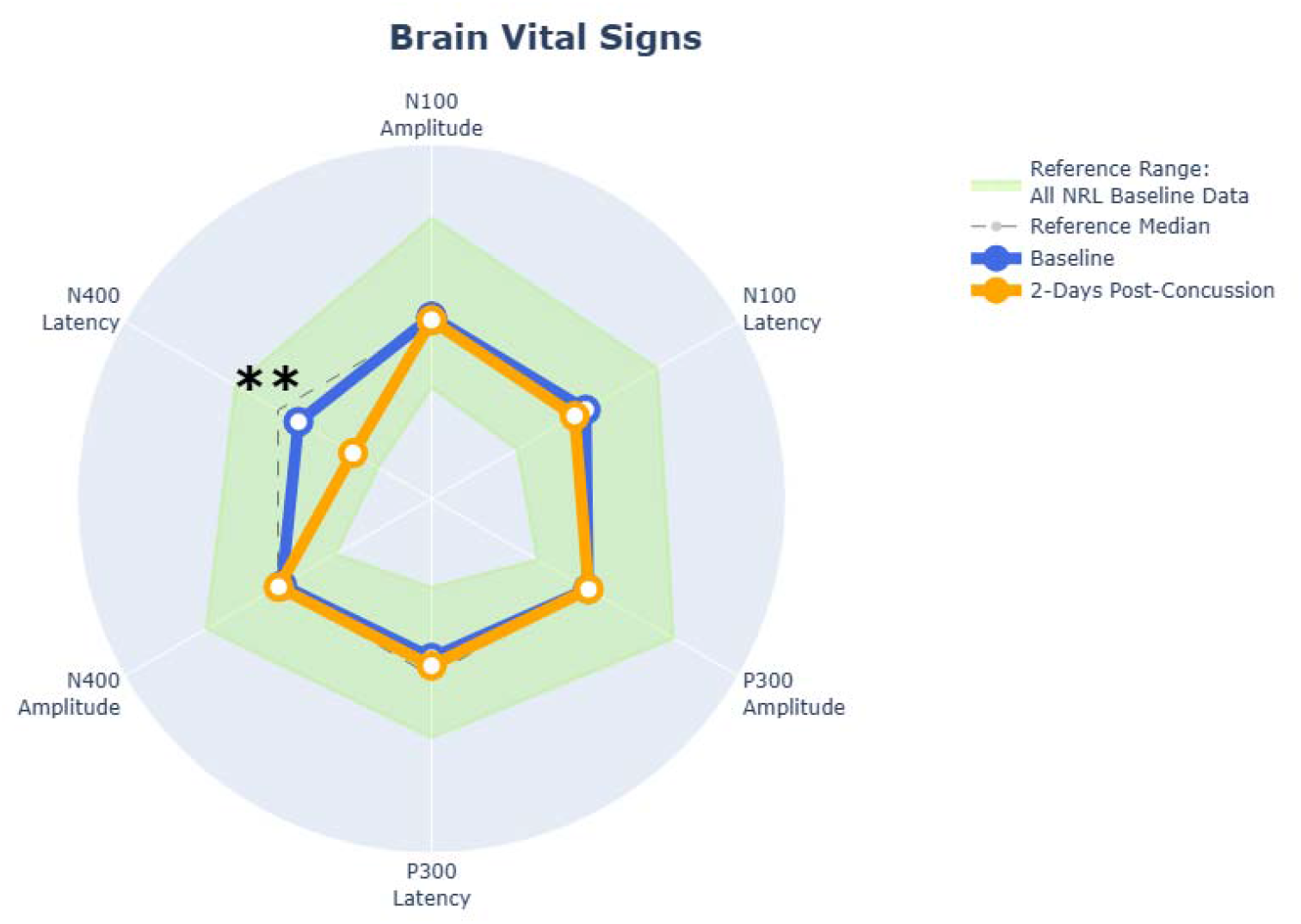
Radar plot overview. Brain vital sign (all six metrics) for the concussed group of athletes. Results compare Baseline and Day 2 time points identifying any concussion related differences. Normative data (green shaded area) shows the athlete specific baseline (*n*=165).

## Results

### Participants

A total of 165 athletes underwent baseline assessment, and 26 athletes (15.8% of the total cohort) sustained a medically diagnosed concussion during the study period. All 26 athletes who sustained a concussion were included in the analysis. Sample characteristics are reported in Table 1. Twenty-one (81%) were male and five (19%) were female athletes. The cohort had a mean age of 22.5 years (SD 3.3; range, 18-33). Most athletes had completed 12 years of education (22/26, 85%) Eighteen players (69%) reported a prior history of at least one concussion. The median number of previous concussions was two (range, 0-5). All 26 athletes returned to play within two weeks, effectively missing one game of rugby league.

**Table 1:** Group demographic characteristics (*n*=26)

| Characteristic | Value |
| --- | --- |
| Age, years mean (SD) | 22.5 (3.3) |
| Age median [min, max] | 22 [18,33] |
| Male n (%) | 21 (80.8) |
| Education, years median [min, max] | 12 [10,19] |
| Any prior concussions n (%) | 18 (69.2) |
| Prior concussions median [min, max] | 2 [0,5] |

Brain vital sign ERP data was obtained for all athletes, except for two athlete time points missing at Day 2 due to a technical error in one athlete and poor data quality in another athlete. By the Day 42 time-point, six athletes were lost to follow up due to non-compliance. The Day 2 time point was carried out at the time of medical follow up, 48 hours after the medical diagnosis of concussion on game day. The Day 5 time point occurred at a mean of 5.7 days (SD 0.84; range, 4-8), the Day 10 time point at a mean of 10.75 days (SD 0.84; range 9-13), and the Day 42 time point at a mean of 43.4 days (SD 0.83; range 5-8).

### Brain vital sign ERPs

All six brain vital sign scores were plotted on standard radar plots to characterise changes across baseline, concussion and return to play time points. Figure 2. outlines changes at Day 2 when compared with baseline. Figure 3. outlines changes across all 5 time points. All LMM timepoint main effect results are provided in Table 2.

**Figure 3:**
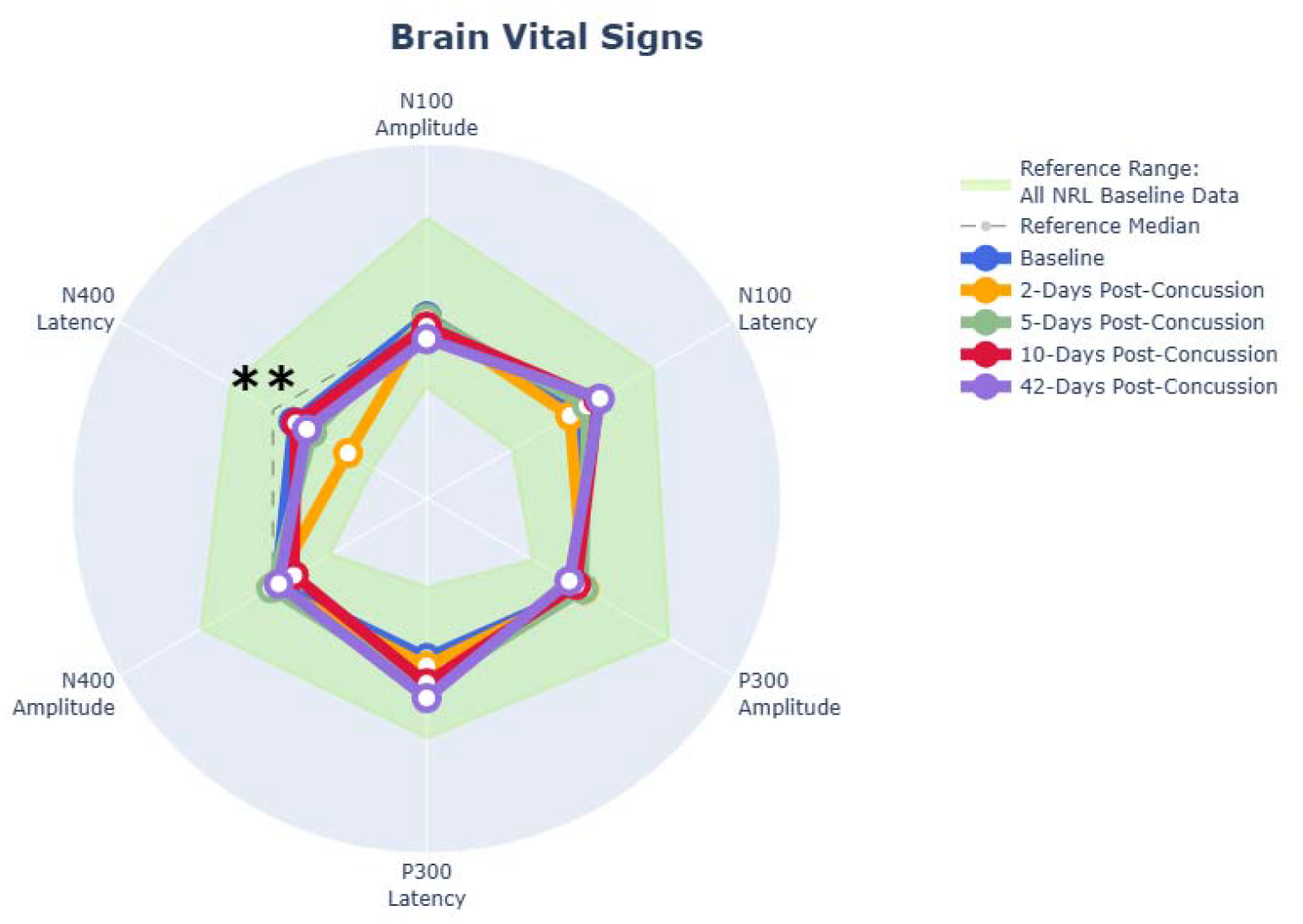
N400 latency changes over time. Group average latencies across all time points with standard error bars. Statistically significant differences between Baseline and Day 2 along with Day 2 and Day 10 are denoted (** *p* < 0.01).

**Table 2:** Brain Vital Sign LMM Results.

| ERP measure | F-Value | p-value |
| --- | --- | --- |
| N100 amplitude | 0.7654 | 0.551 |
| N100 latency | 2.9045 | 0.026 |
| P300 amplitude | 0.8155 | 0.519 |
| P300 latency | 2.8048 | 0.029 |
| N400 amplitude | 1.0521 | 0.385 |
| N400 latency | 4.753 | 0.002 |

#### N400 latency

Figure 3 shows group average data, highlighting N400 concussion related differences in peak latencies. The LMM showed a significant main effect of timepoint (F[4, 91.870] = 4.753, *p* = 0.002). This was further highlighted by two significant post hoc pairwise comparison differences. The first was a significant difference at Day 2 post-concussion compared to baseline (Mean difference = 69.01ms, *p* = 0.002, 95% CI [18, 120.13]). The second was a return of the N400 latency from Day 2 to Day 10 post-concussion (Mean difference = - 66.51ms, *p* = 0.005, 95% CI [14, 119.33]). (Fig. 4). Raw N400 latency timepoint means are included in Supplementary Materials (Supplementary Table 1).

**Figure 4.**
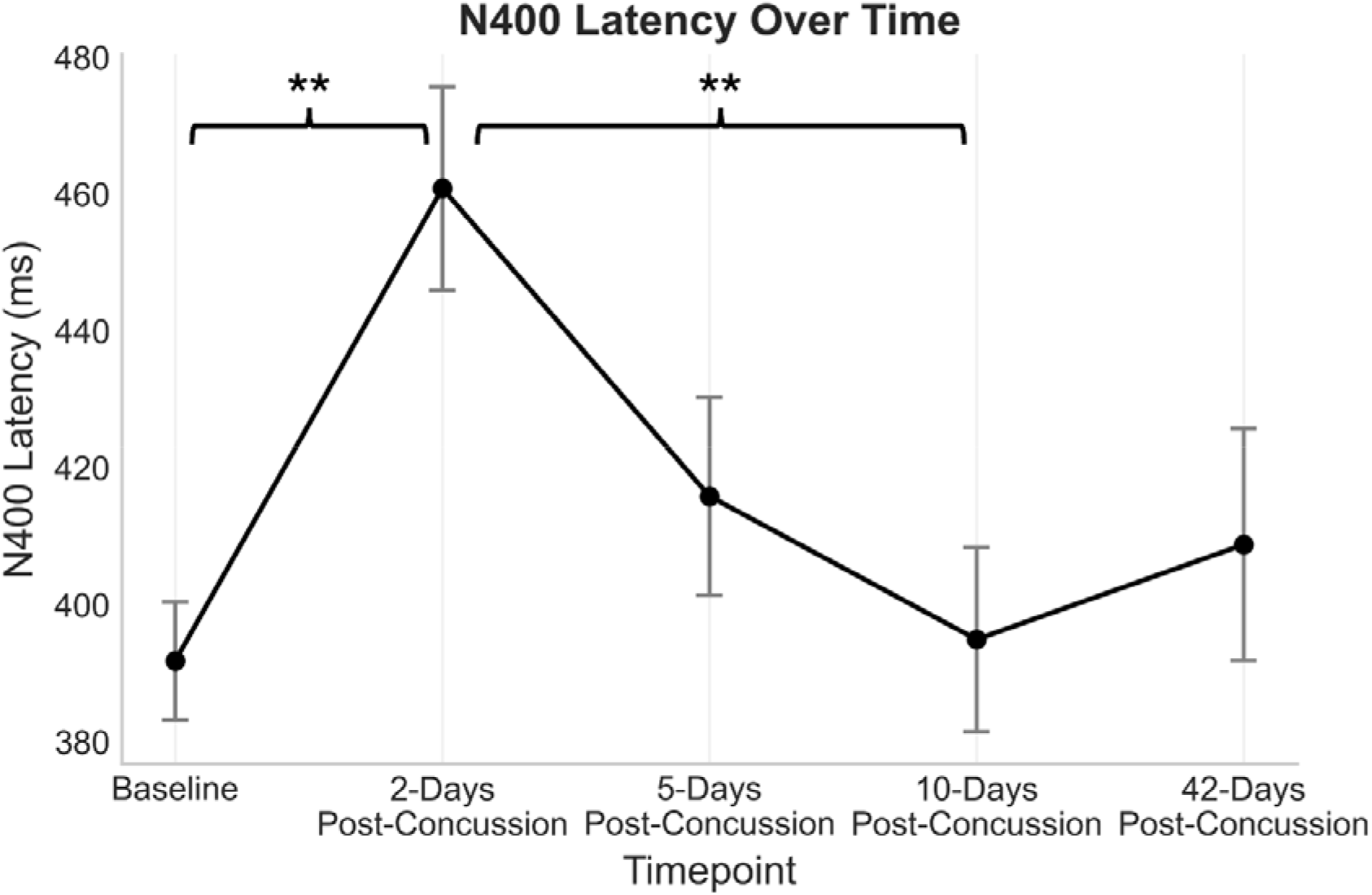
N400 latency individual variance plots. Variance plots demonstrating N400 latency values in each concussed athlete across all timepoints with standard error bars. Individual athlete trajectories are in blue. Statistically significant differences between baseline and Day 2 along with Day 10 are denoted (\**p <*0.01).

Figure 5 provides individual variance across all five time points. Of note, the majority of athlete profiles were consistent with the average profile. One player had a large delay in their N400 latency at the Day 5 timepoint, with increased dispersion towards Day 42. Overall, individuals followed a similar trajectory of a delayed N400 at Day 2, followed by recovery by Day 10.

**Figure 5.**
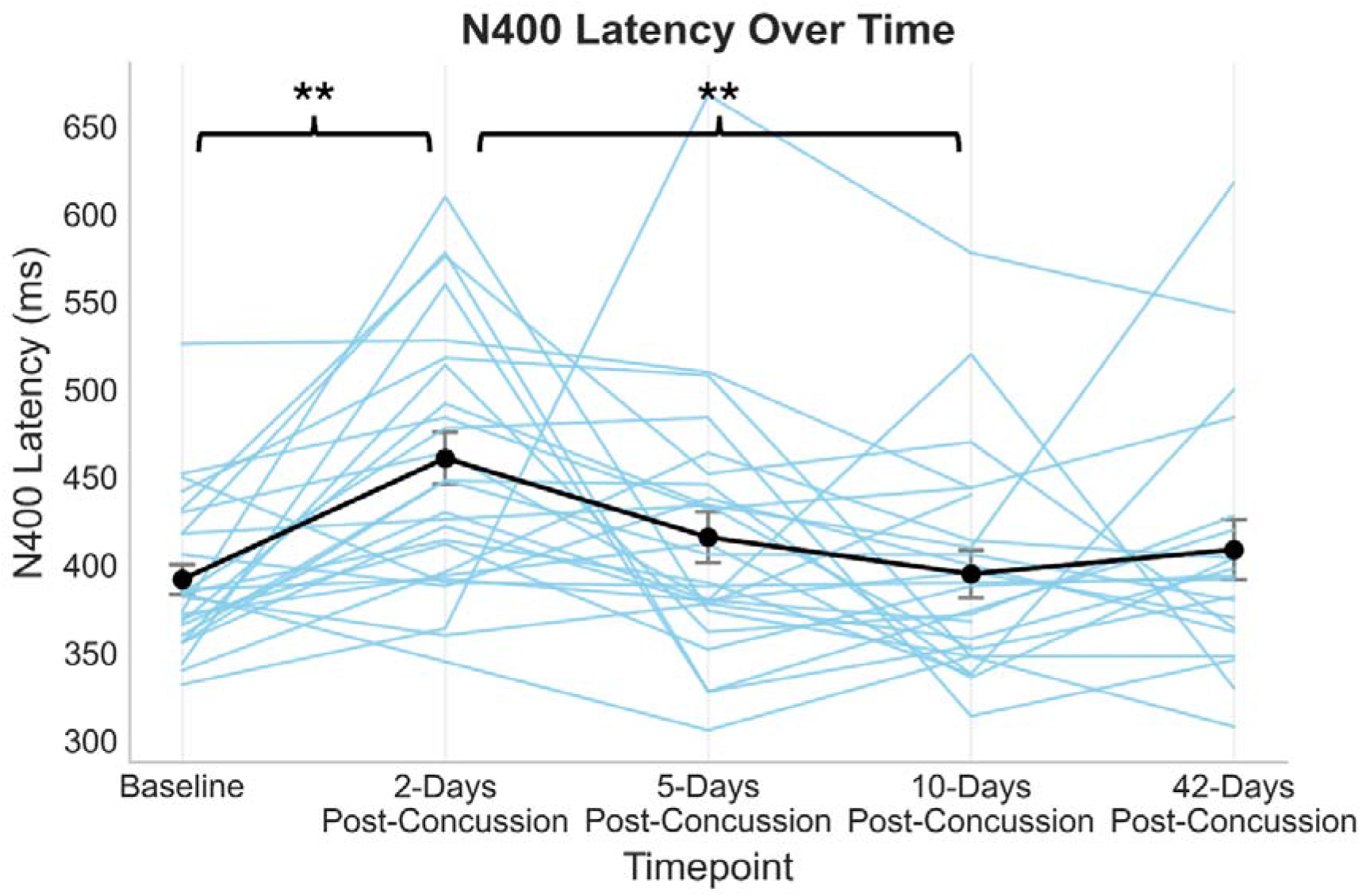
P300 latency over time. Group average latencies across all timepoints with standard error bars. Statistically significant differences between baseline and Day 42 is denoted (\**p <*0.05).

#### N100 and P300

With respect to the N100 and P300 responses, the LMM showed a significant main effect of timepoint for N100 latency (F[4, 90.291] = 2.905, *p* = 0.026) and P300 latency (F[4, 90.200] = 2.805, *p* = 0.029). In post-hoc comparisons, the only significant pairwise comparison across all timepoints for N100 latency and P300 latency that survived adjustment was between baseline and Day 42 post-concussion for P300 latency (Mean difference = 21ms, *p* = 0.0471, 95% CI [0.15, 41.42]). (Fig. 6).

**Figure 6.**
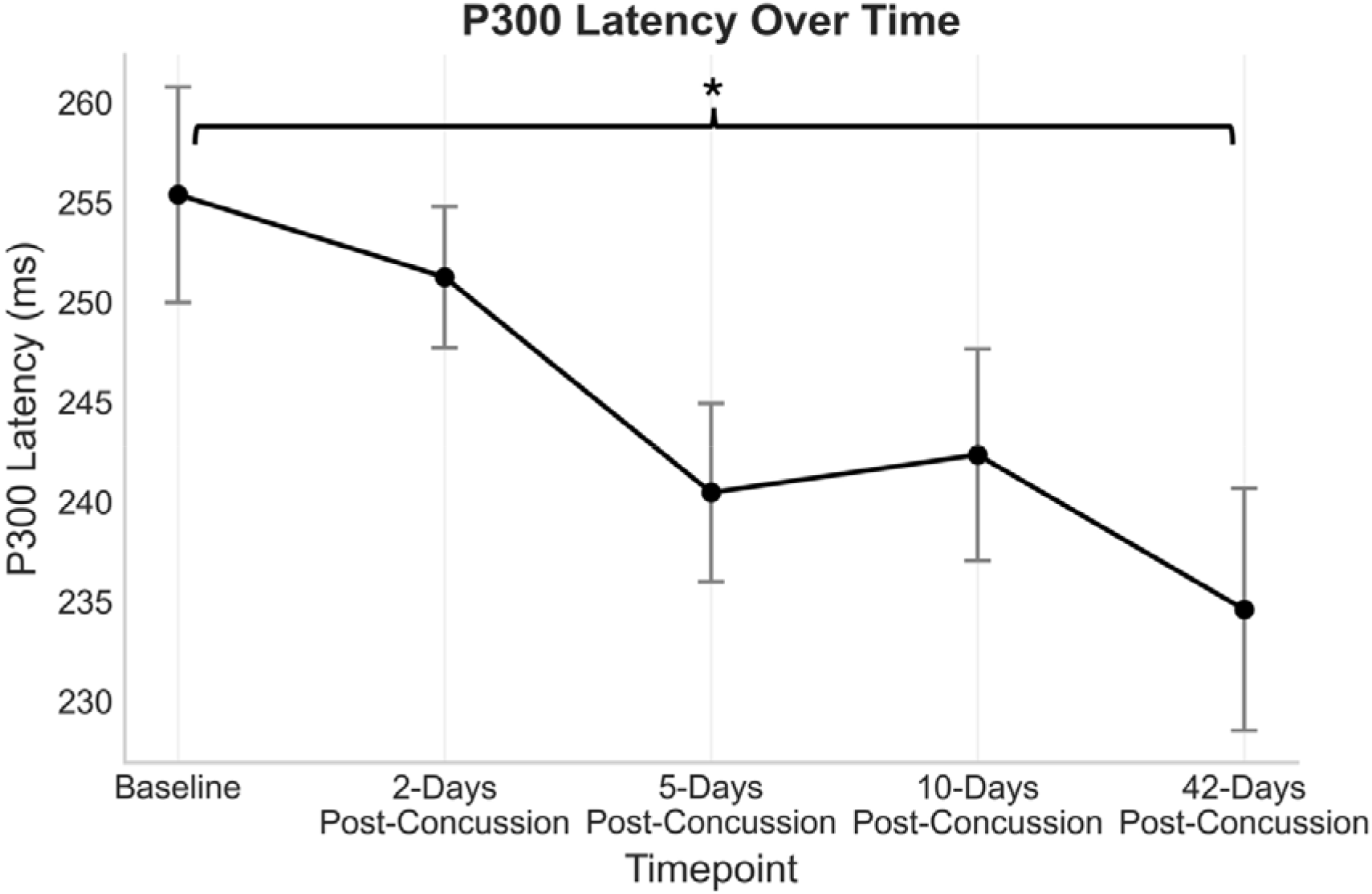
P300 latency over time. Group average latencies across all timepoints with standard error bars. Statistically significant differences between baseline and Day 42 is denoted (\**p <*0.05).

## Discussion

The characterisation of the trajectory of change in the N400 ERP represents an advance in understanding of post-concussion brain neurophysiology and informs growing efforts to integrate biomarkers into concussion diagnosis and recovery. To our knowledge, the current study represents the first demonstration of brain vital sign ERP changes for elite rugby league athlete across the course of their clinical recovery following concussion.

The findings supported the primary hypothesis that objective neurophysiological measures would be sensitive to concussion and would normalise to baseline levels as the athlete recovered from their concussive episode. Notably, the findings revealed an approximately 69 ms average delay in N400 peak latency two days following concussion (p=0.002). This appears to be consistent with prior work in acute concussion in elite ice hockey athletes where N400 delays of around 100 ms of average delay were evident^24^. The N400 latency returned to baseline levels by Day 10 and was noted to return to baseline levels within the NRL’s mandatory 11 day stand-down period.

The specificity of the N400 latency effects represents an interesting finding. While earlier findings in elite ice hockey athletes reported significant changes across all six brain vital sign metrics, these differences were noted in the early acute phase (<24 hours) with a smaller sample size.^24^ The current findings suggest that dynamic changes can be expected on serial ERP assessment in the early recovery phase and differences may be detectable even between the 24 hour and 48 hour mark after concussion. This discrepancy is likely to reflect the underlying metabolic and neurophysiological changes occurring day to day immediately after traumatic brain injury. The isolated latency changes as noted in this study seem to be theoretically consistent with a temporary delay in signal transmission as would be expected with mild axonal injury as opposed to a more diffuse axonal injury resulting in either reduced axonal recruitment, manifesting as a reduced N400 amplitude, or an increased N400 amplitude indicating a compensatory increase in neuronal recruitment in the setting of injury.

The P300 and N100 findings in this study were discordant with those of Fickling *et al.*^24^ where increased N100 and P300 amplitudes were noted in the early acute phase of recovery (< 24hrs) after concussion. Not dissimilar to the N400 findings, these discrepancies may reflect dynamic physiological change that can be expected across the acute, subacute and chronic time periods following concussion. It is interesting to note that an increased P300 amplitude was noted within 24 hours in the ice hockey cohort, values were comparable to baseline values at the 48 hour mark in the current cohort, while previous literature has reported decreased P300 amplitude in the months and years following concussion.^32^ There was a decrease in P300 latency values noted across the timepoints through to the day 42 time point and this may be indicative of a practice effect. Future studies examining additional time points months to years after an event, including year to year scans are likely to shed further light into the directional and quantitative changes that can be expected both acutely and in the longer term.

### Caveats

There are several limitations to this study. The first relates to the sample size of concussed athletes. Despite the sample size in this study being reflective of most published ERP studies, confirmation of our findings in larger sample sizes are needed to further inform the potential utility of cognitive ERPs in assessing concussion recovery. In addition to the monitoring of acute recovery as was undertaken in this study, tracking ERP characteristics at specific, defined subacute and chronic timeframes following concussion would help understand the full spectrum of recovery, particularly in those presenting with persisting symptoms.

This study did not include a comparison group of athletes exposed to RHIs throughout the course of a game but not subsequently diagnosed with concussion. The inclusion of a comparison group and serial assessment of this cohort would help understand the effects of RHIs on cognitive function and objective neurophysiology given growing evidence in the literature around neurophysiological changes, even in the absence of concussion.^25,27,28,33,34^ The findings of a slight increase and dispersion in the N400 latency at Day 42, after returning to play, raises the possibility of RHIs having at least a short term impact on one’s neurophysiology, despite the findings being statistically non-significant. Future studies ought to explore additional time points including year on year changes, to determine whether these changes persist long term.

An interesting finding from the D’Arcy *et al.*^33^ study was a methodological comparison between female and male athletes. The current study investigated both male and female rugby league athletes, however limited sample size for female athletes restricted these analyses. Larger scale studies will assist in determining whether there are group differences between the male and female athlete, especially in the setting where female athletes have been reported to report more pronounced and prolonged symptoms after concussion.^35,36^

## Clinical Relevance and Future Directions

The primary clinical relevance relates to the future potential for incorporation of objective neurophysiological measures into clinical evaluation, as clinical decision assistance. Our findings support the potential role of ERPs and by extension, brain vital signs, as an adjunct to current clinically based judgement on brain recovery post-concussion. In the present study, N400 changes were confined to the acute recovery phase with peak latency returning to baseline levels within a two-week time frame.

Should brain vital sign ERPs be incorporated in the prognostic paradigm for concussion, future research will be required in determining the clinical relevance of persistent ERP abnormalities in the absence of persisting symptoms. It remains unclear whether persistent neurophysiological changes translate into adverse clinical outcomes or are associated with an increased risk of neurocognitive sequelae attributed to repeated head impacts.^7,9^

Furthermore, this study has demonstrated individual variability in N400 ‘normalization’ patterns following concussion. This is likely to be reflective of not only the heterogenous nature of concussion but also the variability in recovery profiles that can be expected following concussion. This raises the possibility of individualized and tailored treatment based off an individual’s N400 recovery profile.

### Conclusion

Brain vital sign ERPs, in particular the N400 latency, is sensitive to the effects of concussion in professional rugby league players with 65-68 ms delayed peak latencies noted 48 hours after concussion. The N400 latency changes returned to baseline levels by Day 10, suggesting neurophysiological recovery in this cohort of professional rugby league players.

## Data availability

The data supporting the findings of this study are available from the corresponding author upon reasonable request. Due to ethical and privacy restrictions related to patient data, the raw data are not publicly available.

## Acknowledgements

We would like to thank the athletes for participating in this study. We acknowledge Mr Craig Catterick, Ms Lisa Annand, Peter Cassar and Kieren Morgan for their input and support in assisting in data collection and Mr Andrew Sargeant for his assistance in interpreting Neurocatch**^®^** scans.

## Supplementary material

**Supplementary Table 1:** Raw N400 latency values for scans.

|  | Baseline | 2-days post-concussion | 5 days post-concussion | 10 days post concussion/prior to RTP | 42 days post-concussion |
| --- | --- | --- | --- | --- | --- |
| Number of scans | 26 | 24 | 26 | 23 | 20 |
| N400 Latency mean and standard deviation (ms) | 392 [43] ms | 461 [71] ms | 416 [72] ms | 395 [63] ms | 409 [74] ms |

## References

1. Broglio SP, Katz BP, Zhao S, McCrea M, McAllister T. Test-Retest Reliability and Interpretation of Common Concussion Assessment Tools: Findings from the NCAA-DoD CARE Consortium. Sports Med. 2018;48(5):1255–1268. doi:10.1007/s40279-017-0813-0

2. Putukian M, Raftery M, Guskiewicz K, et al. Onfield assessment of concussion in the adult athlete. Br J Sports Med. 2013;47(5):285–288. doi:10.1136/bjsports-2013-092158

3. Echemendia RJ, Burma JS, Bruce JM, et al. Acute evaluation of sport-related concussion and implications for the Sport Concussion Assessment Tool (SCAT6) for adults, adolescents and children: a systematic review. Br J Sports Med. 2023;57(11):722–735. doi:10.1136/bjsports-2022-106661

4. Haste P, de Almeida E Bueno L, Jérusalem A, Bergmann J. Performance of current tools used for on-the-day assessment and diagnosis of mild traumatic brain injury in sport: a systematic review. BMJ Open Sport Exerc Med. 2025;11(1):e001904. doi:10.1136/bmjsem-2024-001904

5. Jeter CB, Hergenroeder GW, Hylin MJ, Redell JB, Moore AN, Dash PK. Biomarkers for the diagnosis and prognosis of mild traumatic brain injury/concussion. J Neurotrauma. 2013;30(8):657–670. doi:10.1089/neu.2012.2439

6. Kamil R, Atef AbdelAlim Y, Patel S, et al. Objective Markers for Diagnosing Concussions: Beyond Blood Biomarkers and the Role of Real-Time Diagnostic Tools. J Clin Med. 2025;14(21):7727. doi:10.3390/jcm14217727

7. Patricios JS, Schneider KJ, Dvorak J, et al. Consensus statement on concussion in sport: the 6th International Conference on Concussion in Sport–Amsterdam, October 2022. Br J Sports Med. 2023;57(11):695–711. doi:10.1136/bjsports-2023-106898

8. Smith AM, Stuart MJ, Roberts WO, et al. Concussion in Ice Hockey: Current Gaps and Future Directions in an Objective Diagnosis. Clin J Sport Med. 2017;27(5):503–509. doi:10.1097/JSM.0000000000000412

9. Tabor JB, Brett BL, Nelson L, et al. Role of biomarkers and emerging technologies in defining and assessing neurobiological recovery after sport-related concussion: a systematic review. Published online June 1, 2023. doi:10.1136/bjsports-2022-106680

10. Longworth T, McDonald A, Cunningham C, Khan H, Fitzpatrick J. Do rugby league players under-report concussion symptoms? A cross-sectional study of elite teams based in Australia. BMJ Open Sport Exerc Med. 2021;7(1):e000860. doi:10.1136/bmjsem-2020-000860

11. McLeod S, West SW, Dane K, et al. Disclosure of possible concussions in National Rugby League Women’s Premiership players. J Sci Med Sport. 2025;28(9):707–711. doi:10.1016/j.jsams.2025.04.002

12. NRL Operations Manual. Published online February 2026. Unpublished

13. Echemendia RJ, Brett BL, Broglio S, et al. Sport concussion assessment tool^TM^ - 6 (SCAT6). Br J Sports Med. 2023;57(11):622–631. doi:10.1136/bjsports-2023-107036

14. Collie A, Maruff P, Makdissi M, McCrory P, McStephen M, Darby D. CogSport: reliability and correlation with conventional cognitive tests used in postconcussion medical evaluations. Clin J Sport Med. 2003;13(1):28–32. doi:10.1097/00042752-200301000-00006

15. Prock M, O’Sullivan DM, Tiernan S. Comparing return to play protocols after sports-related concussion among international sporting organisations. Phys Sportsmed. 2024;52(5):421–431. doi:10.1080/00913847.2024.2344432

16. Haider MN, Leddy JJ, Pavlesen S, et al. A Systematic Review of Criteria Used to Define Recovery from Sport-Related Concussion in Youth Athletes. Br J Sports Med. 2018;52(18):1179–1190. doi:10.1136/bjsports-2016-096551

17. Wellm D, Zentgraf K. Diagnostic tools for return-to-play decisions in sports-related concussion. Journal of Concussion. 2023;7:20597002231183234. doi:10.1177/20597002231183234

18. Dockree PM, Robertson IH. Electrophysiological markers of cognitive deficits in traumatic brain injury: a review. Int J Psychophysiol. 2011;82(1):53–60. doi:10.1016/j.ijpsycho.2011.01.004

19. Duncan CC, Summers AC, Perla EJ, Coburn KL, Mirsky AF. Evaluation of traumatic brain injury: brain potentials in diagnosis, function, and prognosis. Int J Psychophysiol. 2011;82(1):24–40. doi:10.1016/j.ijpsycho.2011.02.013

20. Ghosh Hajra S, Liu CC, Song X, et al. Developing Brain Vital Signs: Initial Framework for Monitoring Brain Function Changes Over Time. Front Neurosci. 2016;10:211. doi:10.3389/fnins.2016.00211

21. Ghosh Hajra S, Liu CC, Song X, Fickling SD, Cheung TPL, D’Arcy RCN. Multimodal characterization of the semantic N400 response within a rapid evaluation brain vital sign framework. J Transl Med. 2018;16(1):151. doi:10.1186/s12967-018-1527-2

22. Pawlowski GM, Ghosh-Hajra S, Fickling SD, et al. Brain Vital Signs: Expanding From the Auditory to Visual Modality. Front Neurosci. 2018;12:968. doi:10.3389/fnins.2018.00968

23. Medvedev G, Kirby ED, Harding K, et al. Initial Comparison of Routine Electroencephalography and Event-Related Potentials in Patients with Cognitive Complaints. Can J Neurol Sci. Published online July 24, 2025:1–5. doi:10.1017/cjn.2025.10362

24. Fickling SD, Smith AM, Pawlowski G, et al. Brain vital signs detect concussion-related neurophysiological impairments in ice hockey. Brain. 2019;142(2):255–262. doi:10.1093/brain/awy317

25. Neill MG, Fletcher EKS, Larson E, Emery CA, Smirl JD. Association between downhill mountain bike racing and neurophysiological indices of sensory, attentional and cognitive processing in youth athletes. JSAMS Plus. 2026;8:100152. doi:10.1016/j.jsampl.2026.100152

26. D’Arcy RCN, McCarthy D, Harrison D, et al. An objective neurophysiological study of subconcussion in female and male high school student athletes. Sci Rep. 2024;14(1):28929. doi:10.1038/s41598-024-80262-z

27. Fickling SD, Smith AM, Stuart MJ, et al. Subconcussive brain vital signs changes predict head-impact exposure in ice hockey players. Brain Commun. 2021;3(2):fcab019. doi:10.1093/braincomms/fcab019

28. Fickling SD, Poel DN, Dorman JC, D’Arcy RCN, Munce TA. Subconcussive changes in youth football players: objective evidence using brain vital signs and instrumented accelerometers. Brain Commun. 2022;4(2):fcab286. doi:10.1093/braincomms/fcab286

29. Davis H, Davis PA, Loomis AL, Harvey EN, Hobart G. Electrical reactions of the human brain to auditory stimulation during sleep. Journal of Neurophysiology. 1939;2(6):500–514. doi:10.1152/jn.1939.2.6.500

30. Sutton S, Tueting P, Zubin J, John ER. Information delivery and the sensory evoked potential. Science. 1967;155(3768):1436–1439. doi:10.1126/science.155.3768.1436

31. Kutas M, Federmeier KD. Thirty years and counting: finding meaning in the N400 component of the event-related brain potential (ERP). Annu Rev Psychol. 2011;62:621–647. doi:10.1146/annurev.psych.093008.131123

32. Brush CJ, Ehmann PJ, Olson RL, Bixby WR, Alderman BL. Do sport-related concussions result in long-term cognitive impairment? A review of event-related potential research. Int J Psychophysiol. 2018;132(Pt A):124–134. doi:10.1016/j.ijpsycho.2017.10.006

33. D’Arcy RCN, McCarthy D, Harrison D, et al. An objective neurophysiological study of subconcussion in female and male high school student athletes. Sci Rep. 2024;14:28929. doi:10.1038/s41598-024-80262-z

34. Munce TA, Fickling SD, Nijjer SR, Poel DN, D’Arcy RCN. Mixed martial arts athletes demonstrate different brain vital sign profiles compared to matched controls at baseline. Front Neurol. 2024;15. doi:10.3389/fneur.2024.1438368

35. McGroarty NK, Brown SM, Mulcahey MK. Sport-Related Concussion in Female Athletes: A Systematic Review. Orthop J Sports Med. 2020;8(7):2325967120932306. doi:10.1177/2325967120932306

36. Fahr J, Kraff O, Deuschl C, Dodel R. Concussion in Female Athletes of Contact Sports: A Scoping Review. Orthop J Sports Med. 2024;12(10):23259671241276447. doi:10.1177/23259671241276447

